# Microsimulation based quantitative analysis of COVID-19 management strategies

**DOI:** 10.1101/2021.06.20.21259214

**Authors:** István Z. Reguly, Dávid Csercsik, János Juhász, Kálmán Tornai, Zsófia Bujtár, Gergely Horváth, Bence Keömley-Horváth, Tamás Kós, György Cserey, Kristóf Iván, Sándor Pongor, Gábor Szederkényi, Gergely Röst, Attila Csikász-Nagy

## Abstract

**Background:** Pandemic management includes a variety of control measures, such as social distancing, testing/quarantining and vaccination applied to a population where the virus is circulating. The COVID-19 (SARS-CoV-2) pandemic is mitigated by several non-pharmaceutical interventions, but it is hard to predict which of these regulations are the most effective for a given population.

**Methods:** We developed a computationally effective and scalable, agent-based microsimulation framework. This unified framework was fitted to realistic data to enable us to test control measures (closures, quarantining, testing, vaccination) in multiple infection waves caused by the spread of a new virus variant in a city-sized societal environment. Our framework is capable of simulating nine billion agent-steps per minute, allowing us to model interactions in populations with up to 90 million individuals.

**Findings:** We show that vaccination strategies prioritising occupational risk groups minimise the number of infections but allow higher mortality while prioritising vulnerable groups minimises mortality but implies increased infection rate. We also found that intensive vaccination and non-pharmaceutical interventions can substantially suppress the spread of the virus, while low levels of vaccination and premature reopening may easily revert the epidemic to an uncontrolled state.

**Interpretation:** Our analysis highlights that while vaccination protects the elderly from COVID-19, a large percentage of children will contract and spread the virus, and we also show the benefits and limitations of various quarantine and testing scenarios.

**Funding:** This work was carried out within the framework of the Hungarian National Development, Research, and Innovation (NKFIH) Fund 2020-2.1.1-ED-2020-00003.

**Research in context:** *Evidence before this study:* We still do not have an effective medical treatment against COVID-19 (SARS-CoV-2), thus the majority of the efforts to stop the pandemic focuses on non-pharmaceutical interventions. Each country came up with a local solution to stop the spread of the virus by increased testing, quarantining, lock-down of various events and institutions or early vaccination. There is no clear way how these interventions can be compared, and it is especially challenging to predict how combinations of interventions could influence the pandemic. Various mathematical modelling approaches helped decision-makers to foresee the effects of their decisions. Most of these models rely on classical, deterministic compartmental “SEIR” models, which can be solved easily but cannot take into account spatial effects and most non-pharmaceutical interventions affect the same parameters, so there is no way to analyse their separate or joint effects. Agent-based microsimulations are harder to solve but can consider far more details. Several models were developed on these lines focusing on questions about ideal vaccination, lock-down or other specific problems, but none of these studies evaluated and compared the individual and mixed effects of a wide variety of control measures.

*Added-value of this study:* Here we present PanSim, a framework where we introduce a detailed infection event simulation step and the possibility to control specific workplaces individually (schools, hospitals, etc.), test various vaccination, testing and quarantine scenarios while considering preconditions, age, sex, residence and workplace of individuals and mutant viruses with various infectivity. The level of details and granularity of simulations allow our work to evaluate this wide range of scenarios and control measures accurately and directly compare them with one another to provide quantitative evidence to support decision-makers. Analysis of our simulations also provides emergent results on the risks children and non-vaccinated individuals face.

*Implications of all the available evidence:* The agent-based microsimulation framework allows us to evaluate the risk and possible consequences of particular interventions precisely. Due to the outstanding efficiency of the computations, it is possible to apply scenario-based analysis and control design methods which require a high number of simulation runs to obtain results on a given confidence level. This will enable us to design and quantitatively assess control measures in case of new waves of COVID-19 or new pandemic outbreaks.

## Introduction

Epidemic management includes a variety of control measures ranging from non-pharmaceutical interventions such as social distancing, testing and quarantining^1,2^, to vaccination^3^, hospitalisation, and beyond^4–7^. Typically, control measures are differentially applied to various groups (compartments) of the society and decision-makers often need to refocus their intervention strategies as new infection hotspots or new virus variants emerge^8,9^. Mathematical modelling is now increasingly used to inform decision-makers^10,11^. In these models, disease progression is often described by variants of the now classical SEIR approach where individuals move between disease-related compartments (such as susceptible, exposed, infected, and removed^12^). A key element is the probability of person-to-person disease transmission upon adequate contact, which defines the likelihood of a person’s transition from the susceptible to the exposed compartment. This is often based on predefined values, but it can also be calculated from environmental data, using formulas of varying complexity^13–15^. Traditionally, the resulting models are then run with ordinary differential equations, which are deterministic and can provide crude estimates on how interventions may affect the outcome of the epidemic. Differential equations can be relatively easily fitted to pandemic data^16–20^ but have difficulty dealing with population heterogeneity and spatial context at sufficient resolution, especially as intervention strategies often change. For instance, differential equation driven analysis of vaccination strategies or other intervention policies is feasible and widely used^19,20^, but the number of compartments is limited and predominantly organised by (only a couple of parameters, e.g.) age or serostatus. Another strategy is provided by stochastic, agent-based models (ABMs), where agents, corresponding to individuals, move and transmit the infection among each other. ABMs can easily handle subgroups, complex scenarios and also provide indications regarding the geographic spread of the pandemics. However, they are compute-intensive as they rely on repetitions of many elementary steps, and also, a large amount of external data is required for their parameterisation^5,21–24^. There is a clear need for compute-efficient fine-grained ABM implementations to support decision-makers in planning and scheduling effective interventions such as vaccination policies and safe reopening schemes.

## Methods

Here we present a modelling approach using the notions of control theory wherein a detailed, agent-based, microsimulation description was built for a mid-sized Hungarian town using realistic statistics on the population as well as on its daily movements. We used this Pandemics Simulator model (for brevity *PanSim*) to simulate the COVID-19 (SARS-CoV-2) pandemics starting from the onset of the second wave in the Autumn of 2020 and continuing through the Spring of 2021 until September 2021. During this time, various lock-down and opening measures were implemented, a new, fast-spreading virus variant appeared, and vaccination programs were started. All these events affected the dynamics of the pandemic and have been incorporated into the model for detailed analysis. A special focus of the simulations was the design of vaccination and reopening strategies used to inform decision-makers responsible for practical implementations.

Although the presented results are focused on a single town, PanSim can be applied to larger populations, and in fact, our results were used to assist Hungarian decision-makers in designing control measures. The performance of the core simulator enables the expansion of the model to a far larger population sizes. The limiting factor is the availability of uniformised data on population movements, but data from mobile network providers and services like Google Maps could overcome this barrier.

### Modelling Framework

In our framework, agents represent people who live in a virtual city and follow daily routines consisting of regular activities such as going to work or school, resting during weekends, as well as stochastically selected elements such as shopping, entertainment, etc. Such an ensemble of agents is a typical complex, stochastic system^25^ where agents pass infection randomly to each other, and pandemic is a state of the system with an infection level above a threshold. Interventions, such as lock-downs, quarantines target specific parts of the system intending to bring the system back to a “healthy” regime best described as an unrestricted state with no or low infection levels. This setup is analogous to controlling dynamical systems, a problem that occurs in many fields^26^. From this perspective, an intervention is a control measure (input) characterised by a few parameters (constraints), such as target (scope), speed, and resource requirements (costs). For instance, a lock-down measure can target a geographic region, or an age-group, or a type of business (say restaurants) or a given time of the day. A particular feature of pandemic management is that a series of interventions or scenarios are applied. For example, vaccination programs or reopening plans are complex scenarios wherein simple interventions are carried out according to a given schedule in time and space. Precisely, in the presented micro-level model, 179,500 agents follow the statistical behaviour of the population of the Hungarian town of Szeged. At the heart of the model is an infection event, where virus transmission occurs with a probability depending on the proportion of infectious individuals present at a given location such as a home, classroom, or hospital ward^22^. A fast-spreading virus variant – mastered on the example of B.1.1.7 that appeared in late 2020^27,28^ – was considered to increase transmission probability by a factor of 1·5 to 1·9^29,30^. Disease progression was considered to follow a SEIRD-like model^9^ in which an individual can be in a susceptible, exposed, infected, recovered, or deceased state (see supplement for details). Vaccination was included in the model as a reduction in individuals’ probability of contracting the virus. It was reduced to 52% and 96% after 12- and 28-days post-vaccination^31^, respectively. Before the simulation, PanSim was provided with input data such as the number, age-, sex-distribution, medical precondition, etc., of the agents, the structure of families assigned to various geographic locations, size of classes at schools etc. (see supplement for details). At the beginning of the simulation, PanSim randomly assigned the agents to various locations (homes, schools, working places, etc.) as well as to daily routines (e.g., home-work-shopping-home). The agents were then let to follow their routine in the microsimulation (Fig. 1A), and they passed on the infection to each other with some probability, which depends on their infection status. In the one-year-long simulations, starting from 1st October 2020, additional lock-down and curfew restrictions were implemented on 11th November 2020, and vaccination was started on 1st January 2021 (see supplement for detailed parameter sets). The B.1.1.7 variant was introduced into the population starting 11th January, leading to the third wave of infections in the Spring of 2021.

**Figure 1:**
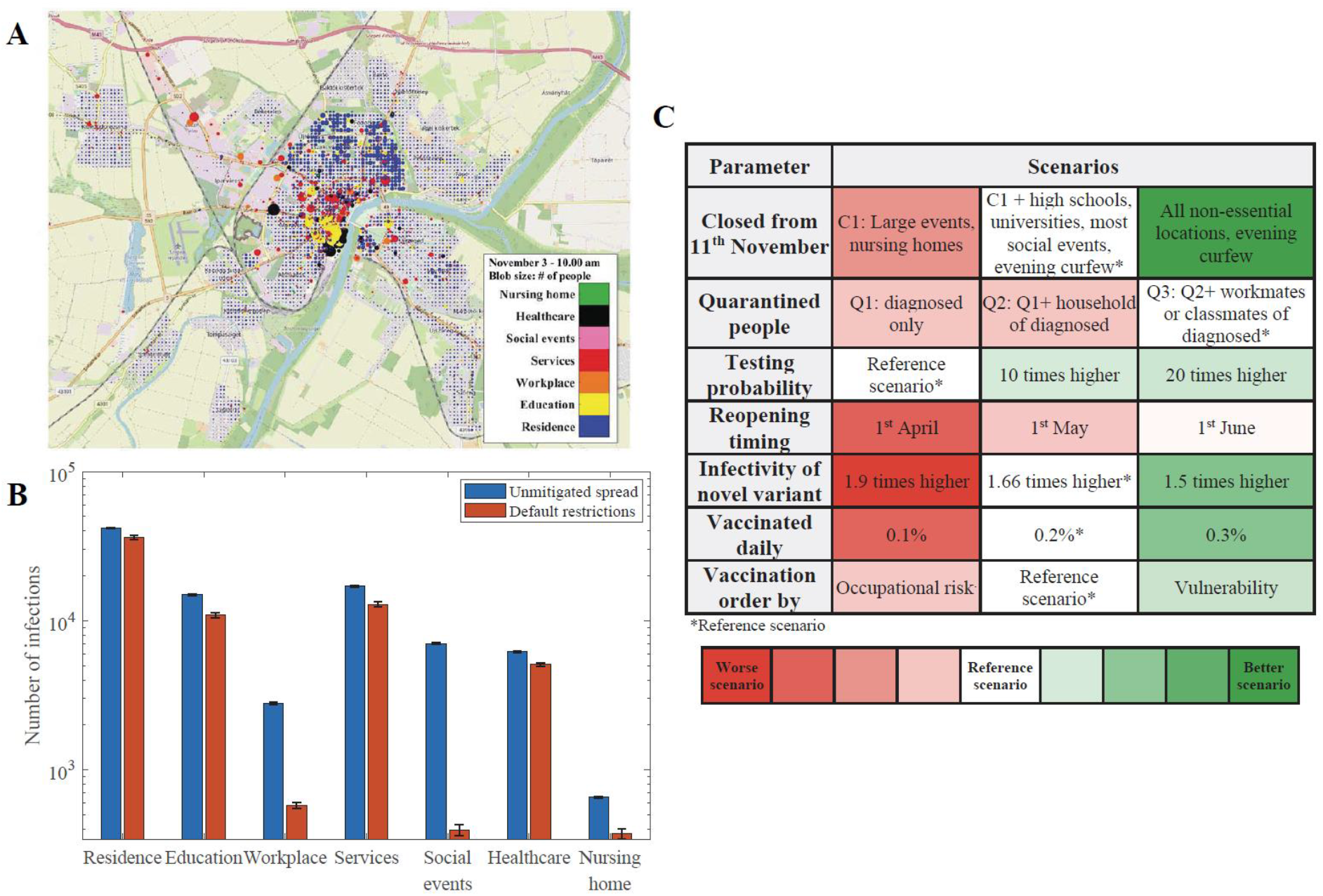
PanSim realistically simulates population movements and infections in the virtual city emulating Szeged. (A) Institutions and households are localised in a virtual city. Colour codes represent different types of locations, and node sizes correlate with the number of infected people. (B) Distribution of infection events at various types of locations when the virus is unmitigated or when restrictions are applied. (Note the logarithmic scale, error bars show the uncertainty of 20 simulations.) (C) Sensitivity of the pandemic in various intervention types and levels. The colour code shows the severity function of the pandemics for the labelled changes. (Details on the analysed scenarios and the severity function can be found in the supplement.)

## Results

Simulation outputs give geographic information about the number of simulated people at each location (Fig. 1A) and statistics on infection events at these locations (Fig. 1B). It is noticeable that restrictions decrease the number of infections generally, but the significant effects concentrate on institute types that are locked down during these restrictions (Fig. 1B). Closing particular types of locations is just one of many policies applied to control the pandemic. The modelling framework is flexible since interaction strategies (quarantine policies, closures of specific locations, testing intensities, and reopening timings) can be implemented by simply changing input parameters rather than reprogramming the system. This makes it possible to calculate „intervention landscapes” in which these interventions and other key parameters (infectivity of a novel strain, vaccination intensities, and prioritisation order) are varied in a grid-like fashion, and the outcome of a given scenario is characterised by a colour code (Fig. 1C). A “severity function” was calculated for the measure of the seriousness of the pandemia by adding the total number of deaths to a scaled function of the total number of hospital beds occupied above a critical limit for the whole investigated period. This analysis shows that stronger interventions (such as high vaccination rates, stringent restrictive measures), as expected, tend to improve outcomes while other factors, for instance, the infectivity of the virus variants or too early reopening, can markedly deteriorate the outcome (Fig. 1C). From these strategies, we will now concentrate on a few key control parameters of the pandemic that plague decision-makers in many countries.

### Quarantine strategies can have mixed effects with the arrival of a fast-spreading and more harmful virus variant

Quarantine procedures can follow different rules. If only the diagnosed infected patients are quarantined (Q1), and there is no contact tracing for quarantining, then the autumn wave is much larger, and most of the population get through the disease to reach herd immunity before the fast-spreading virus variant appears (Fig. 2A). With better tracking, resulting in quarantining households (Q2) as well as the classmates or a portion of workmates (Q3), the autumn wave is smaller, leaving more people susceptible for the more harmful variant in the spring wave (Fig. 2A). The severity of the pandemic is still much higher with a weaker quarantine policy because the hospital burden is exceptionally high for an extended period during the autumn wave (Fig. 1C, Fig. 2A). The simulator suggests that the eventual total number of deaths is higher with better quarantining policies, partly because a more transmissible and harmful variant appeared later (Fig. 2B). However, death rates would be higher when healthcare is overloaded, which is not accounted for in the simulator. We have also assumed that people who got infected by the slower spreading variant are fully immune to the new variant as well after recovery^32^.

**Figure 2:**
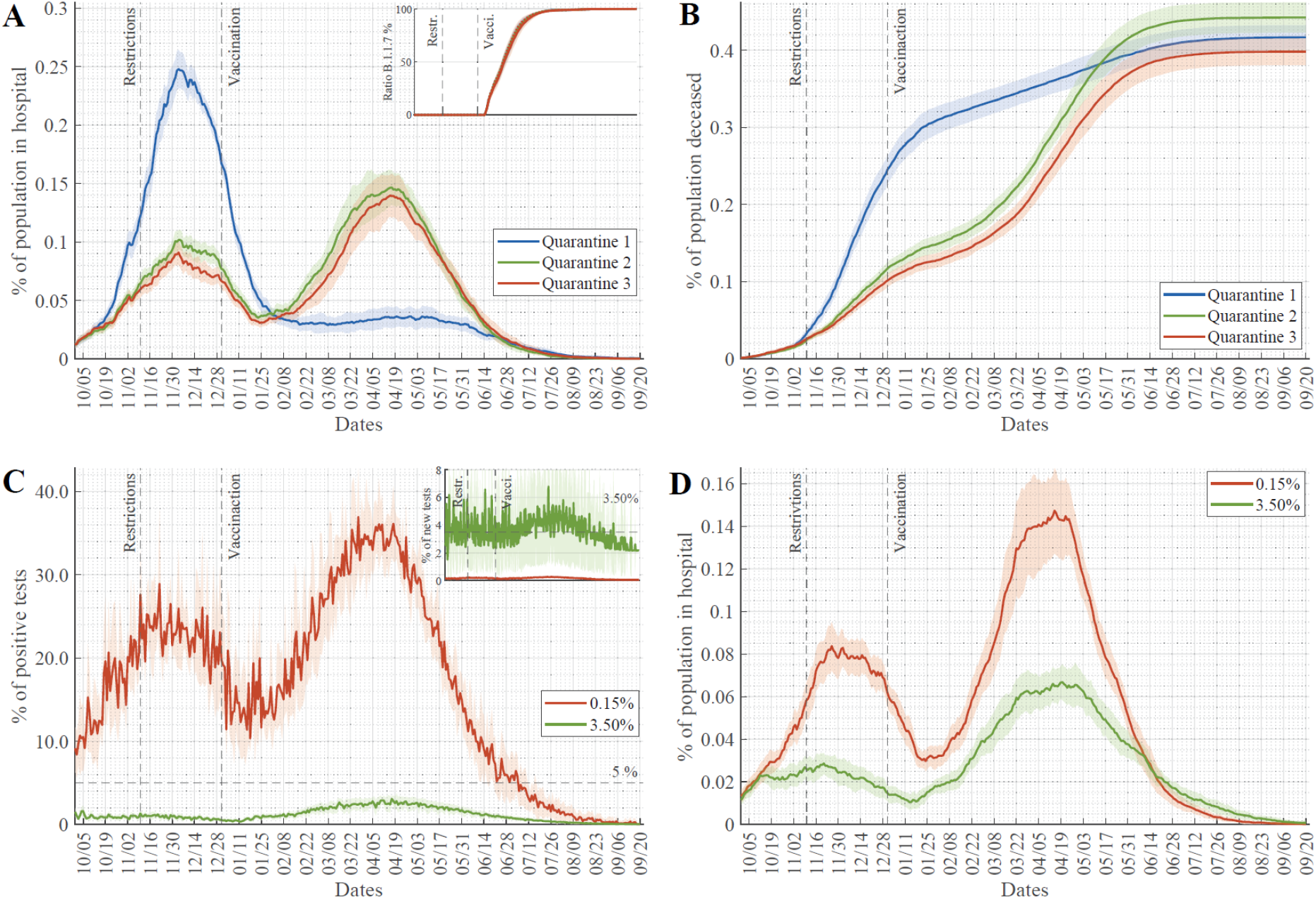
Variations in quarantine scenarios and testing intensities can slow down but not suppress the pandemics. (A-B) Time course of simulations with varied quarantine scenarios (Q1 – diagnosed patient quarantined at home, Q2 – diagnosed patient and household quarantined, Q3 – patient, household and workmates/classmates are quarantined) showing (A) the percentage of hospitalised COVID-19 patients in the population and (B) the accumulated number of death events due to COVID-19 scaled to the whole population. Appearance and spread of the B.1.1.7 variant are shown on the inset of panel A. Start date of restrictions and the start of vaccination are noted on plots. (C-D) The testing rate was increased from the reference value of 0.15% of people tested daily, fitted to Hungarian data (Supplementary Figure 4), to the highest level achieved by most actively testing countries (3·5% daily). (C) Time courses of the daily ratio of positive tests and (D) the percentage of hospitalised COVID-19 patients in the population each day (hospital burden) are plotted. Daily testing rates are shown on the inset of panel C. (Average and std. of 20 simulations are shown.)

### Increased testing intensity can effectively suppress only moderately infective virus strains

The WHO recommends that the ratio of positive tests should remain under 5% in order to control pandemics. With our simulations fitted to the Hungarian testing rate (Supplementary Figure 4) (testing ∼0·15% of the population daily), we see far higher positivity values (Fig. 2C). If we increase the testing rate to ∼3·50% of the population tested each day (see supplement for details) – which level was reached only by a few countries – then we can push the positivity rate below 5% (Fig. 2C). The model follows here the “contact tracing” concept, where housemates, class-/workmates of positively tested individuals are tested the next day with some probability (these rates were increased in simulations on Fig. 2 C,D). With testing, ∼3·50% of the population each day, the positivity rate is pushed below 3% (Fig. 2C), but even with such low positivity, we find a severe hospital burden during the spring wave (Fig. 2D). We can conclude that extensive testing could repress the pandemic with a moderately infective virus (average European strains spreading during the autumn wave) (Fig. 2D). However, extensive testing is incapable of controlling the pandemic of a highly transmissible and more harmful variant, such as B.1.1.7 (spring wave of Fig. 2D), despite vaccination starting around the same time as the appearance of this variant (Fig. 2A inset).

### Vaccination order strategies have opposing effects on new infection numbers and hospital burden

Ever since the COVID-19 vaccination started towards the end of 2020, vaccines were in short supply in many countries, so decision-makers had to design vaccination strategies that determined whom to vaccinate and in what order (e.g., differences between Israel and the EU)^33^. One approach is to concentrate on vulnerable groups, such as the elderly, the chronically ill, etc. These groups were then vaccinated in the order of decreasing vulnerability. Another approach is to select groups according to their essentiality and occupational risk, such as healthcare professionals, nursing home workers, teaching personnel, etc. Both ordering methods could have their benefits, and various countries follow either one of these or a mixed prioritisation rule. In earlier plots, we assumed a mixed order used in Hungary. Still, we have also analysed the differences caused by using the two extreme strategies (see supplement for details of different vaccination orders). The occupational risk method is better in controlling infection numbers and helps to maintain key infrastructure (e.g. essential workers), but the number of the hospitalised and deceased people is lower in the case of vulnerability-based prioritisation (Fig 3 A-C). The reason for this difference is apparently that the elderly and people with chronic conditions who otherwise make up the majority of hospitalised and deceased patients are vaccinated first, and so they avoid infection. Early vaccination of highly vulnerable groups can in itself reduce the hospital burden even at higher infection rates in the spring wave (Fig 3B). Nevertheless, the key strategy to control the pandemic is to increase the vaccination intensity (Fig. 3D), which could have a far larger effect than varying the vaccination order (compare Fig. 3B – before reopening and Fig. 3D).

**Figure 3:**
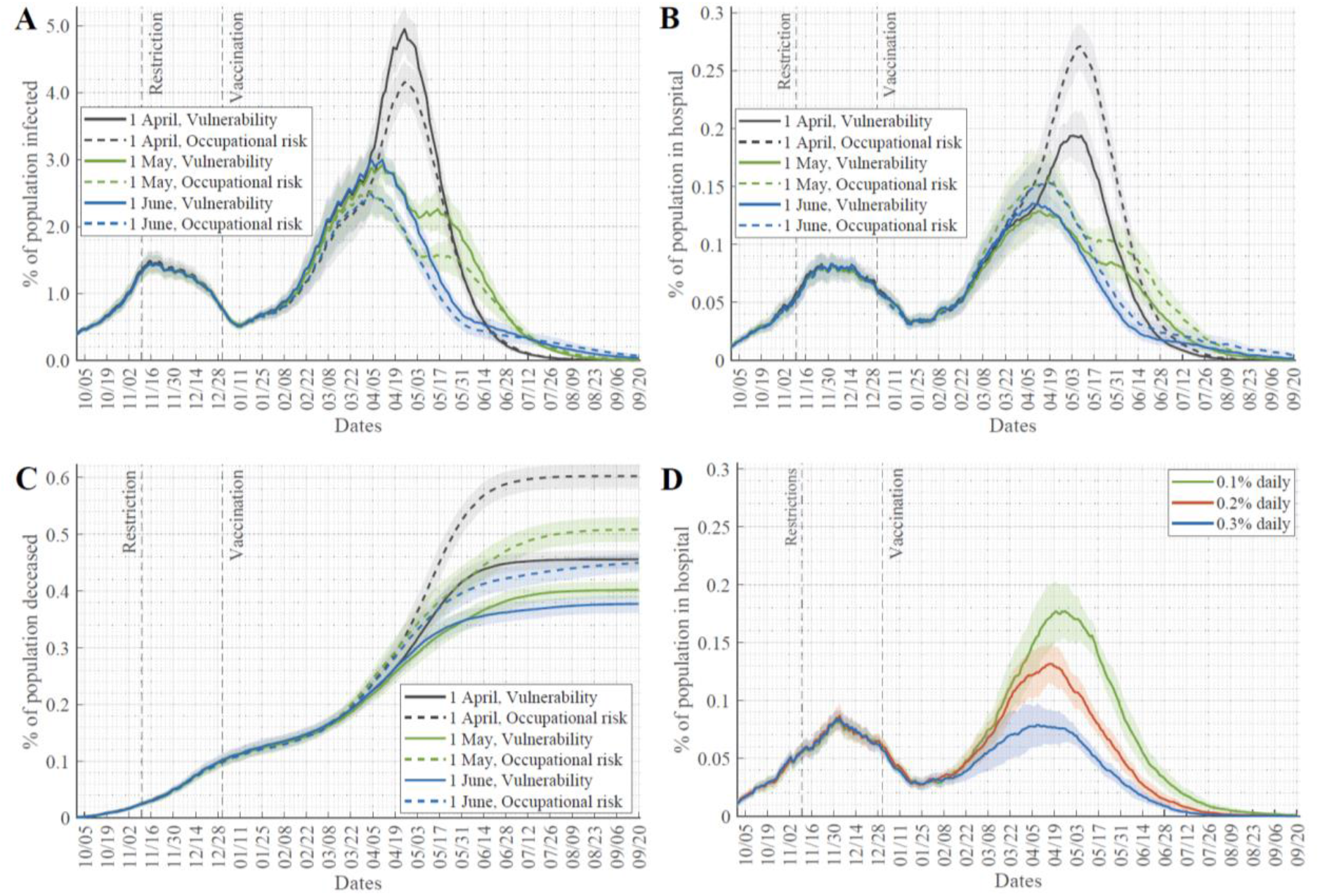
Results of various vaccination and reopening strategies. (A) Percentage of the population infected, (B) percentage of hospitalised COVID-19 patients in the population, (C) the accumulated number of death events due to COVID-19 scaled to the whole population, each day. Panel A-C were simulated with the assumption that daily 0.2% of the population could be vaccinated. (D) Changes in this rate have major effects on the percentage of hospital patients. (Average and std. of 20 simulations are shown.)

### Too early reopening after lock-downs can lengthen pandemics

Curfew in evening hours and lock-down of social event sites, restaurants, and pubs is a crucial strategy to control pandemics by decreasing the number of interactions between individuals. In the simulations presented, all of these restrictions were applied from November 2020 (dashed line in plots of Fig. 2 and 3). A crucial question is when these restrictions can be lifted, and life can return to “normal”.

In Figure 3 A-C, we also analysed the effects of lifting these restrictions at three different time points. Reopening close to the peak of the pandemic can lead to the collapse of the healthcare system by filling up hospitals (Fig. 3B) and increase the total death cases (Fig. 3C). Reopening while the number of infections is dropping could extend the length of the pandemic leading to a slight increase in total death counts, but not affect the hospital burden too dramatically (Fig. 3 B, C).

## Discussion

The modelling framework described here was primarily developed to handle lock-down, quarantine, and vaccination scenarios. Nevertheless, relevant new information emerged from the simulation results. For instance, it shows that the autumn wave affected the various age groups quite similarly, while in the larger spring wave, distinct age groups were differentially involved (Fig. 4A). In these simulations, we used the vulnerability vaccination order, resulting in a situation where the elderly are less often infected than younger people in the spring wave. Specifically, almost 50% of children below 14 who meet in schools (students over 14 are home-schooling in these simulations) go through the infection during the spring wave (Fig. 4A).

**Figure 4:**
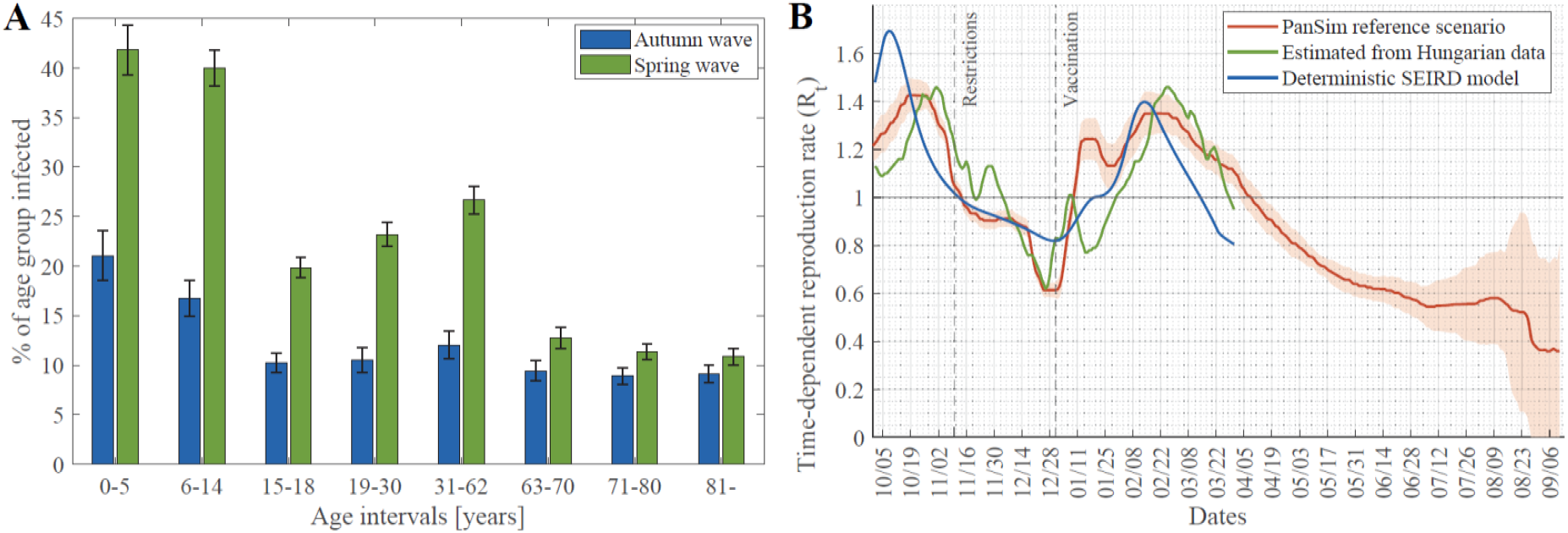
High infection rate of children and precise, effective reproduction number (R_t_) emerge from the simulation results. (A) Percentage of the various age groups catching the infection during the autumn and the spring wave (error bars show the uncertainty of 20 simulations). (B) Changes in the reproduction number of the virus were calculated from the reference scenario (Fig. 1C) simulations (average and std. of 20 simulations) plotted together with the fits of a deterministic SEIRD model and empirical data about Hungary^34^.

We can calculate the critically investigated effective reproduction number (R_t_), defined as the actual average number of secondary cases per primary case at a given time (see supplement for details). When R_t_ is below one, the epidemic is declining, while the number of cases is increasing when it is above one. Plotting the simulated changes of the effective reproduction number in time, together with the same values empirically calculated for Hungary and the solution of a deterministic model^9^, we can observe that PanSim microsimulation provides a more detailed and better prediction of the observed data (Fig. 4B).

PanSim can be applied to complex, realistic scenarios and can test the effects of various interventions. Here we used these features to gain a general understanding of the impact of various quarantine strategies, possibilities to contain the pandemic by increased testing, optimal vaccination ordering and risks of early reopening. Directions of further development include differentiation between vaccine efficacies and taking into consideration the waning of immunity. The emergence of new virus variants with their specific epidemiological parameters and potential capability to escape previously acquired immunity could also be considered in an extended version of the model. From an algorithmic perspective, an intervention in PanSim is a uniformly built GPU (graphics processing unit) compatible module that has input parameters such as the type of intervention, speed, resource availability etc., and that can be combined into realistic scenarios (see https://github.com/khbence/pansim and supplement for details). The model could be adapted to specific data on any other cities, regions, or countries. Simulations of one year considered with a ten-minute timestep for 179,500 agents were run under 64 seconds on a single NVIDIA V100 GPU. Still, the code is scalable up to 90 million agents on a single GPU. This computational speed enables PanSim to be used for control design, where interventions are optimised towards specific goals (e.g., keeping healthcare burden below a threshold while minimising the costs caused by quarantining people)^9,12^. Events with finer spatial resolution, for example, using public transport or working in separated offices, are feasible to implement in case of sufficient input data. The performance can also be scaled up to country or continent level simulations, including occasional long-distance trips (e.g. tourism or business) that could be crucial in the importation of diseases. The major challenge here is collecting the data on individuals’ movements to feed into the simulator. However, with extensive, uniformised data^35^ it will be possible to use PanSim to study detailed and dynamically changing country-specific intervention strategies in fine spatio-temporal resolution upon this, or possible later pandemics.

## Data Availability

The code used for the simulations is available here: https://github.com/khbence/pansim, and it contains links to the data used to run the scenarios described.

## Acknowledgements

We thank colleagues at the Lechner Knowledge Center, GeoX Kft. and at Szeged Municipality for data sources.

## Author Contributions

IZR and BKH developed the tool, IZR, DC, JJ, ZB, GH, and TK designed and performed simulations, KT, GH, TK and GR collected data, KT built the database, all authors analysed and interpreted results, SP, GS, GR and ACN supervised the project, IZR, JJ, SP, ZB, KT and ACN wrote the paper by contributions to all authors, IZR, DC, JJ, SP, GS, GR and ACN designed the study. All authors approved the submitted version.

## Additional Information

Supplementary Information is available for this paper. Correspondence and requests for materials should be addressed to IZR or ACN.

## References

1 Peak CM, Kahn R, Grad YH, et al. Individual quarantine versus active monitoring of contacts for the mitigation of COVID-19: a modelling study. Lancet Infect Dis 2020; 20: 1025–33.

2 Wells CR, Townsend JP, Pandey A, et al. Optimal COVID-19 quarantine and testing strategies. Nat Commun 2021; 12: 1–9.

3 Forni G, Mantovani A, Forni G, et al. COVID-19 vaccines: where we stand and challenges ahead. Cell Death Differ. 2021; 28: 626–39.

4 Ferguson NM, Cummings DAT, Fraser C, Cajka JC, Cooley PC, Burke DS. Strategies for mitigating an influenza pandemic. Nature 2006; 442: 448–52.

5 Chang SL, Harding N, Zachreson C, Cliff OM, Prokopenko M. Modelling transmission and control of the COVID-19 pandemic in Australia. Nat Commun 2020; 11: 1–13.

6 Nadanovsky P, dos Santos APP. Strategies to deal with the COVID-19 pandemic. Braz. Oral Res. 2020; 34. DOI:10.1590/1807-3107BOR-2020.VOL34.0068.

7 Sturniolo S, Waites W, Colbourn T, Manheim D, Panovska-Griffiths J. Testing, tracing and isolation in compartmental models. PLoS Comput Biol 2021; 17: e1008633.

8 Ecdc. SARS-CoV-2 - increased circulation of variants of concern and vaccine rollout in the EU/EEA - 14th update. https://www.ecdc.europa.eu/en/covid-19/timeline-ecdc-response.This (accessed 20th March, 2021).

9 Péni T, Csutak B, Szederkényi G, Röst G. Nonlinear model predictive control with logic constraints for COVID-19 management. Nonlinear Dyn 2020; 102: 1965–86.

10 Vespignani A, Tian H, Dye C, et al. Modelling COVID-19. Nat. Rev. Phys. 2020; 2: 279–81.

11 Perra N. Non-pharmaceutical interventions during the COVID-19 pandemic: A review. Phys. Rep. 2021; published online 13th February. DOI:10.1016/j.physrep.2021.02.001.

12 Röst G, Bartha FA, Bogya N, et al. Early phase of the COVID-19 outbreak in hungary and post-lockdown scenarios. Viruses 2020; 12: 1–30.

13 Watanabe T, Bartrand TA, Weir MH, Omura T, Haas CN. Development of a dose-response model for SARS coronavirus. Risk Anal 2010; 30: 1129–38.

14 Zhang N, Huang H, Su B, Ma X, Li Y. A human behavior integrated hierarchical model of airborne disease transmission in a large city. Build Environ 2018; 127: 211–20.

15 de Oliveira PM, Mesquita LCC, Gkantonas S, Giusti A, Mastorakos E. Evolution of spray and aerosol from respiratory releases: theoretical estimates for insight on viral transmission. Proc R Soc A Math Phys Eng Sci 2021; 477: 20200584.

16 Liu M, Thomadsen R, Yao S. Forecasting the spread of COVID-19 under different reopening strategies. Sci Rep 2020; 10: 1–8.

17 Pinto Neto O, Kennedy DM, Reis JC, et al. Mathematical model of COVID-19 intervention scenarios for São Paulo-Brazil. Nat Commun 2021; 12: 418.

18 Reiner RC, Barber RM, Collins JK, et al. Modeling COVID-19 scenarios for the United States. Nat Med 2021; 27: 94–105.

19 Matrajt L, Eaton J, Leung T, Brown ER. Vaccine optimisation for COVID-19: Who to vaccinate first? Sci Adv 2020; 7: eabf1374.

20 Bubar KM, Reinholt K, Kissler SM, et al. Model-informed COVID-19 vaccine prioritisation strategies by age and serostatus. Science (80-) 2021;: eabe6959.

21 Chang S, Pierson E, Koh PW, et al. Mobility network models of COVID-19 explain inequities and inform reopening. Nature 2021; 589: 82–7.

22 Chinazzi M, Davis JT, Ajelli M, et al. The effect of travel restrictions on the spread of the 2019 novel coronavirus (COVID-19) outbreak. Science (80-) 2020; 368: 395–400.

23 Hoertel N, Blachier M, Blanco C, et al. A stochastic agent-based model of the SARS-CoV-2 epidemic in France. Nat Med 2020; 26: 1417–21.

24 Rockett RJ, Arnott A, Lam C, et al. Revealing COVID-19 transmission in Australia by SARS-CoV-2 genome sequencing and agent-based modeling. Nat Med 2020; 26: 1398–404.

25 Galea S, Riddle M, Kaplan GA. Causal thinking and complex system approaches in epidemiology. Int J Epidemiol 2010; 39: 97–106.

26 Nise NS. Control Systems Engineering, 8th Edition | Wiley. Wilney, 2020.

27 Davies NG, Jarvis CI, van Zandvoort K, et al. Increased mortality in community-tested cases of SARS-CoV-2 lineage B.1.1.7. Nature 2021;: 1–5.

28 Volz E, Mishra S, Chand M, et al. Assessing transmissibility of SARS-CoV-2 lineage B.1.1.7 in England. Nature 2021;: 1–6.

29 Supasa P, Zhou D, Dejnirattisai W, et al. Reduced neutralisation of SARS-CoV-2 B.1.1.7 variant by convalescent and vaccine sera. Cell 2021; 184: 2201-2211.e7.

30 Brookman S, Cook J, Zucherman M, Broughton S, Harman K, Gupta A. Effect of the new SARS-CoV-2 variant B.1.1.7 on children and young people. Lancet Child Adolesc. Heal. 2021; 5: e9–10.

31 Polack FP, Thomas SJ, Kitchin N, et al. Safety and Efficacy of the BNT162b2 mRNA Covid-19 Vaccine. N Engl J Med 2020; 383: 2603–15.

32 Arafkas M, Khosrawipour T, Kocbach P, et al. Current meta-analysis does not support the possibility of COVID-19 reinfections. J Med Virol 2021; 93: 1599–604.

33 Cylus J, Panteli D, van Ginneken E. Who should be vaccinated first? Comparing vaccine prioritisation strategies in Israel and European countries using the Covid-19 Health System Response Monitor. Isr. J. Health Policy Res. 2021; 10: 16.

34 Arroyo-Marioli F, Bullano F, Kucinskas S, Rondón-Moreno C. Tracking R of COVID-19: A new real-time estimation using the Kalman filter. PLoS One 2021; 16: e0244474.

35 Buckee CO, Balsari S, Chan J, et al. Aggregated mobility data could help fight COVID-19. Science.). 2020; 368: 145–6.

