## Supplementary Material for "Microsimulation based quantitative analysis of COVID-19 management strategies"

### Data Integration

The agent-based simulator requires detailed information about simulated individuals (age, daily behaviour) and their visited locations. Due to the sensitive nature of personal and movement information of the city's residents, it was not made available for the experiments. Therefore realistic synthetic input data has been constructed based on census statistical information and geographic databases. This section briefly summarises the data sources and data generation algorithms.

#### Data Sources

The Hungarian Central Statistical Office published freely accessible statistical data on the population, including geographical distribution, age distribution, families' constitution (number of parents, children, grandparents, or senior citizens), and statistical information of households. These data are based on the population censuses (conducted every ten years) and population micro-censuses. Detailed statistics are available on the Csongrád-Csanád County (NUTS Level 3) and Szeged District (NUTS Level 4 / LAU Level 1).

The other significant source of data was the geographic distribution of residential locations of Szeged. The data was made available through the TEIR (information system for development purposes on settlements/areas) of the city municipality. This database provided the points of interest in Szeged that places have been used to assign workplaces and other places to the residents. The capacity (size) of locations that are visited regularly by agents has been estimated manually based on publicly available information.

Geographical data have been extracted from the official land register. The data has been transformed to eliminate sensitive information: The area of Szeged has been partitioned into 100 $m^2$ square-shaped cells containing each cell's statistics (location, number of residents).

The City of Szeged's local administration provided real and current information about public schools (kindergartens, primary and secondary schools). The population of University students has been estimated based on the data available publicly on the University of Szeged web page.

#### Data Synthesis

A greedy algorithm has been implemented to construct a realistic population of the city randomly. The necessary steps are the following:

1. First, the algorithm creates a family then its members. The size and constitution of the family are based on the statistics of the population census (see the right panel of Supplementary Figure 1).
2. Once the family is generated, it is being placed in any of the available cells. This step assigns a residential location to each person of the family. During this step, the artificially created families are placed in real-

world locations where families reside with a similar constitution (see the left panel of Supplementary Figure 1).

3. As primary locations are assigned, the secondary locations are drawn. Depending on the age of a person, a school, university, or workplace will be assigned. (The algorithm primarily seeks a school in the proximity, but workplaces can be located in farther places (see Supplementary Figure 2)).
4. The third round of location assignment adds other places of interest (such as shops, parks, social locations, healthcare-related locations, etc.) that the agents can visit.
5. Additional agents are added, like tourists and commuters.

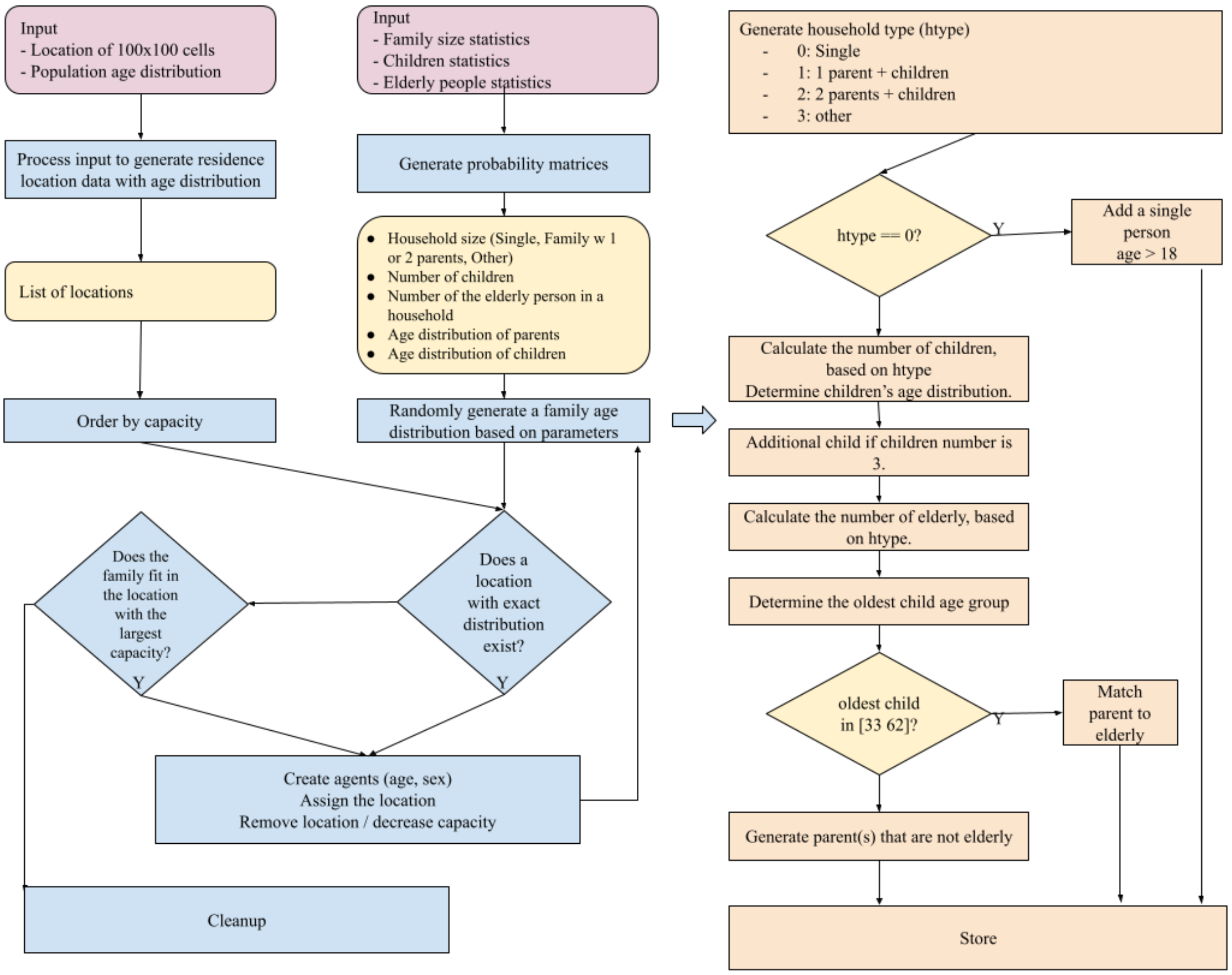


**Supplementary Figure 1:** Algorithm and data flow to generate and initialise households, families and agents.

### Validity check

The realisticness of the generated data is checked with the following steps:

- The family and age distribution (of people generated in step 1) is calculated and matched to the real information. As the population information and statistical information are from different years (2011 and 2016 vs 2018), a slight difference is present. The difference between the number of agents and the actual population is less than 3%.
- The age distribution of the cells (and residential cell assignment) are verified in an independent step. The difference in age distribution is somewhat more enormous (6%-8% depending on the actual generation). The reason is that we had the age distribution of the County and the District instead of the city itself. However, the geographical data is current and contains only the city.

- School assignments are accurate, but workplace assignments are not verified because there was no reliable, publicly available information on that.
- Interesting locations are generated randomly. (Basic places such as groceries, pharmacies, etc., are always assigned, and the simulator makes sure that residents visit such places.) The random assignment of other locations (like pubs, social places) is not unrealistic at all.

Once previous steps are performed, additional visitor agents are generated, such as tourists and commuters (both workers and students). Commuters behave like ordinary agents except that their residence is outside of the city. Statistical information is obtained from the publicly available census data. Tourists are assigned with public locations and hotels as residence. The average number of nights spent in the area is retrieved from the Hungarian Central Statistical Office's statistical databases.

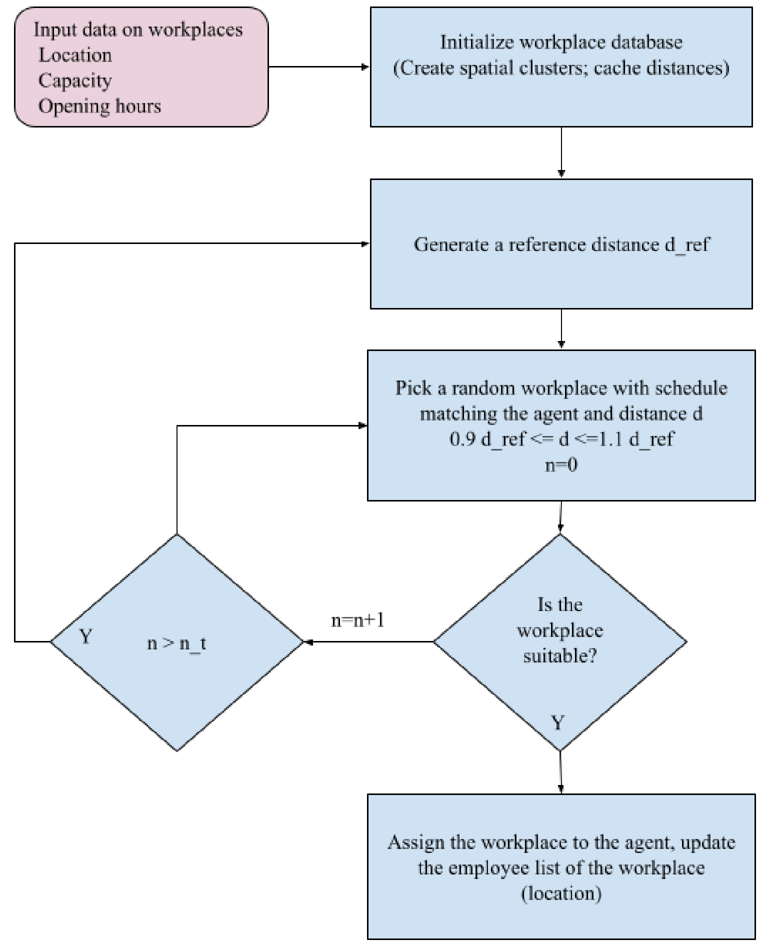


**Supplementary Figure 2:** Algorithm and data flow to assign workplaces for agents, according to their types.

### Movement of agents

Each agent in the model has a distinct type based on its age (e.g. infant or elementary school student) or its profession (e.g. full-time worker, employee with atypical working hours like medical professionals or person who do not have a workplace or who work from home) (Supplementary Table 1). The agents also have a predefined list of places, including their home, school, or workplace, and a set of other locations they can visit (Supplementary Table 2). Categories of the places can be found in Supplementary Table 3.

| | |
|---|---|
| 1 | Infant |
| 2 | Kindergarten student |
| 3 | Elementary school student |
| 4 | High school student |
| 5 | University student |
| 6 | Full-time worker (standard 9-17 schedule) |
| 7 | Afternoon shift worker (12-20 schedule) |
| 8 | Stay-at-home schedule (retired, unemployed, working from home) |

**Supplementary Table 1:** Types of agents. The agents are organised into these categories based on their age and daily routine (lifestyle).

<table>
<tr><th></th><th>Infant</th><th>Kinder-garten student</th><th>Elementary school student</th><th>High school student</th><th>University student</th><th>Full time 9-17 worker</th><th>Afternoon shift worker</th><th>Stay-at-home schedule</th></tr>
<tr><td>Public space</td><td bgcolor="#e77c73"></td><td bgcolor="#e77c73"></td><td bgcolor="#e77c73"></td><td bgcolor="#e77c73"></td><td bgcolor="#e77c73"></td><td bgcolor="#e77c73"></td><td bgcolor="#e77c73"></td><td bgcolor="#e77c73"></td></tr>
<tr><td>Residence</td><td bgcolor="#e77c73"></td><td bgcolor="#e77c73">**</td><td bgcolor="#e77c73">**</td><td bgcolor="#e77c73">**</td><td bgcolor="#e77c73">**</td><td bgcolor="#e77c73">**</td><td bgcolor="#e77c73">**</td><td bgcolor="#e77c73">***</td></tr>
<tr><td>kindergarten, elementary school, high school *</td><td></td><td bgcolor="#e77c73"></td><td bgcolor="#e77c73"></td><td bgcolor="#e77c73"></td><td></td><td></td><td></td><td></td></tr>
<tr><td>School classes</td><td></td><td></td><td bgcolor="#e77c73"></td><td bgcolor="#e77c73"></td><td></td><td></td><td></td><td></td></tr>
<tr><td>University *</td><td></td><td></td><td></td><td></td><td bgcolor="#e77c73"></td><td></td><td></td><td></td></tr>
<tr><td>standard-schedule full time</td><td></td><td></td><td></td><td></td><td></td><td bgcolor="#e77c73"></td><td></td><td></td></tr>
<tr><td>a workplace with non-standard schedule</td><td></td><td></td><td></td><td></td><td></td><td></td><td bgcolor="#e77c73"></td><td></td></tr>
<tr><td>activity site for short (10-30 min) visits *</td><td></td><td></td><td></td><td bgcolor="#e77c73"></td><td bgcolor="#e77c73"></td><td bgcolor="#e77c73"></td><td bgcolor="#e77c73"></td><td bgcolor="#e77c73"></td></tr>
<tr><td>activity site for longer (30-120 min) visits *</td><td></td><td bgcolor="#e77c73"></td><td bgcolor="#e77c73"></td><td bgcolor="#e77c73"></td><td bgcolor="#e77c73"></td><td bgcolor="#e77c73"></td><td bgcolor="#e77c73"></td><td bgcolor="#e77c73"></td></tr>
<tr><td>evening-schedule social activity site for 20-100 people *</td><td></td><td></td><td></td><td></td><td bgcolor="#e77c73"></td><td bgcolor="#e77c73"></td><td bgcolor="#e77c73"></td><td bgcolor="#e77c73"></td></tr>
</table>

| | | | | | | | | |
|---|---|---|---|---|---|---|---|---|
| evening-schedule social activity site for 100-1000 people * | | | | (red) | (red) | (red) | (red) | (red) |
| weekend-schedule social activity site for 50-500 people * | | | | (red) | (red) | (red) | (red) | (red) |
| a daytime recreational site for a long stay | (red) | | | (red) | (red) | (red) | (red) | (red) |
| Health Centre * | (red) | (red) | (red) | (red) | (red) | (red) | (red) | (red) |
| Hospital * | (red) | (red) | (red) | (red) | (red) | (red) | (red) | (red) |

**Supplementary Table 2:** Agent - location pairs. Agents of each agent type can visit those types of locations that are signed with red in the table. Red cell: Agents have that location type on their location list. White cell: Agents normally do not visit that type of location. *workspace for some agents; **the residence of some workers can be outside of the city (commuters); ***the residence for older people can be a nursing home

| Category description | Examples for typical locations |
|---|---|
| Public space | street |
| Residence | households |
| Nursing home | |
| Commuter box | residence of commuters outside the town (not simulated) |
| Daytime stay site for children | kindergarten, elementary school, high school |
| School classes | |
| University | |
| Standard-schedule full-time workplace | office, pharmacy, factory |
| A workplace with a non-standard schedule | afternoon, evening shift |
| Activity site for short (10-30 min) visits | shop, front office, pharmacy |
| Activity site for longer(30-120 min) visits | shop, front office, playground, gym |
| Evening-schedule social activity site for 20-100 people | restaurant, pub |
| Evening-schedule social activity site for 100-1000 people | cinema, theatre, concert hall |
| Weekend-schedule social activity site for 50-500 people | church |
| Daytime recreational site for extended stay (hours) | city park, outdoor |
| Health Centre | |
| Hospital | |

**Supplementary Table 3**: Types of locations. Locations are organised into these categories at the initialisation step of the model.

For example, students arrive at school in the morning gradually within a time interval and spend some of their days in their classroom with a group of other children of the same age and other parts of their school day in a common location with all the other students and school workers (accounting for the breaks and lunchtime between classes). The students can go home or participate in some extracurricular activity (e.g. music, sports, etc.) with a certain probability after school. (The same is true for kindergarten students while infants stay home most of the day.) High school students can additionally visit shops, social events and some services (e.g. theatre, cinema). Full-time workers can go shopping, visit social events and use other services (e.g. restaurants, pubs, gyms, parks) after their working hours or during their lunchtime. Agents with non-standard workplace are also able to use these opportunities when they are not at work, while agents representing people without a workplace (home office, unemployed, retired) can do that all day. University students are similar to full-time workers, but they spend a more variable time interval at the university. Every agent finishes their day at home. (More technical details can be found in the Movement section of Simulator functionalities.)

There are some dynamic factors that influence regular activity schedules. Weekends and holidays have distinct activity patterns. Students and full-time workers are not at school or at work, and they spend more time (with higher probability) with free time activities, social events, or shopping. (Workers with non-standard schedules can work these days.) The general health status, the well-being state of the agents (healthy, light, mild, severe symptoms) is updated daily based on the COVID status (agent type and well-being state-dependent activity patterns can be found at https://github.com/khbence/pansim). It also influences the movement patterns. Agents with light symptoms have a higher chance to skip free time activities and stay home. Mild symptoms increase the chance of absence from school or from work and staying home or visiting a health centre or a hospital (where the infection is automatically diagnosed). Severe cases get into the hospital, and they become inactive until they get better (or die). Quarantine forces the agents to stay home. Closure of some places (e.g. social events) prohibits them from visiting specifically these locations while introducing increased awareness of the epidemics generally decreases the probability of going out and increases the time spent at home after work or school or at the weekends.

Each time frame (e.g. arriving at the workplace, arriving home, doing shopping) is represented as an interval in the model in order to avoid unrealistic synchronised movements of the agents but still express daily periodicity (e.g. rush hours). The weekend-weekday periodicity was parameterised to fit with real-life mobility data[1–3]. The decrease of general outgoing was estimated based on Facebook's Movement Range Map. Other time intervals and the probability profiles of visiting certain places were constructed in a way to mimic the behaviour of a typical Hungarian urban population qualitatively. The detailed agent type, date, and well-being dependent activity patterns are available at [https://github.com/khbence/pansim].

### Integration of a SEIRD model

An extended SEIRD model describes the progression of the COVID-19 epidemic in the simulation environment based on the works of Röst et al.[4] and Péni et al.[5] This model assigns SARS-COV-2 infection-related states to the members of the agent population: they are susceptible (S), exposed to the disease (E), infected (I), recovered and immune (R) or dead (D). The infected state is divided further into subcategories depending on the severity of the symptoms from presymptomatic to critical (hospitalisation in intensive care unit) states. Recovered and recovering states are also distinguished, and the latter refers to not infectious patients who still need medical help in a hospital in their recovery after a critical condition. Each state has typical length, infectiousness, assigned well-being states (Supplementary Table 4), and progression opportunities (Supplementary Figure 3).

| State code | State name | Average days in the state | Maximum number of days in the state | Infectiousness (Inf) | Well-being state |
|---|---|---|---|---|---|
| S | Susceptible | - | - | 0 | Healthy |
| E | Exposed | 3.2 | 9 | 0 | Healthy |
| I1 | Presymptomatic | 2 | 5 | 1 | Healthy |
| I2 | Asymptomatic | 3 | 10 | 0.75 | Healthy |
| I3 | Light symptomatic | 3 | 10 | 1 | Light |
| I4 | Mild symptomatic | 3 | 10 | 1 | Mild |
| I5H | Severe symptoms (hospitalised) | 10 | 20 | 0.1 | Severe |
| I6H | Critical (hospitalised in intensive care) | 10 | 20 | 0.1 | Severe |
| RH | Recovering (from critical hospitalised) | 14 | 21 | 0 | Severe |
| R | Recovered / Resistant | 365 | 730 | 0 | Healthy |
| D | Deceased | - | - | - | Deceased |

**Supplementary Table 4:** Disease transmission states. The table summarises the parameters of disease transmission states.

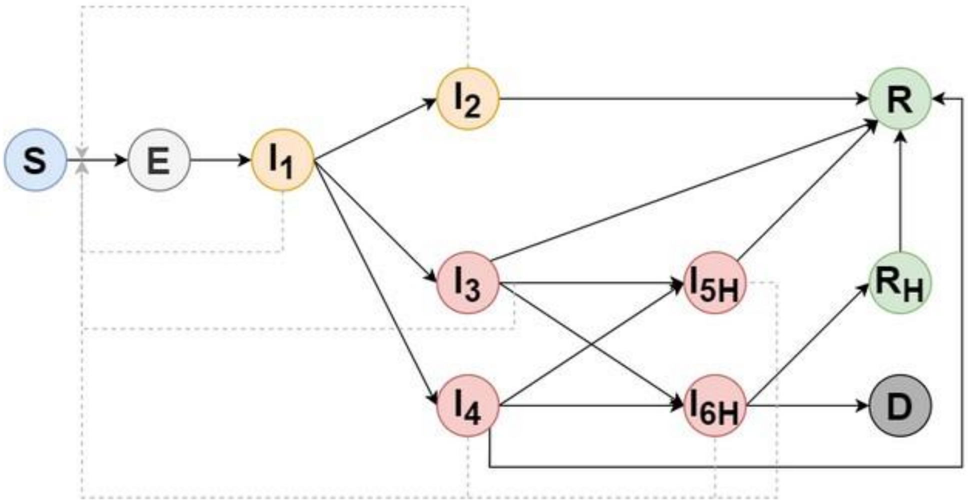


**Supplementary Figure 3:** COVID-19 disease transmission state graph. The disease progresses based on an extended SEIRD model. It is a modified version of the model of Röst et al.[4], and Péni et al.[5] Solid lines denote the possible state transitions, and dotted lines represent the infection (start from the infectious states (I) and end at susceptible (S) - exposed (E) transition). Orange nodes correspond to no symptoms, red nodes to worse progressions. Green nodes mean the non-infectious, recovering/recovered states (R), and agents in states signed with "H" are in hospital. The worst progression goes towards the deceased state (D). More details about the properties of each state can be found in Supplementary Table 4.

The disease transmission model published by Röst et al.[4] was modified based on the following points:

- the symptomatic state is divided into subgroups with different well-being parameter: there are agents with light and mild symptoms (I3, I4)
- patients in the hospital can infect
- the duration in a given state has a geometric distribution

We used data also from another publication (Péni et al.[5]) to estimate relative infectiousness and average lengths of given states.
Susceptible agents can get infected and enter the exposed state by spending time in a location shared with the infectious agent(s). The probability of an individual becoming infected primarily depends on the ratio of infectious individuals in the location. Further details about the infection model are discussed in the Infection dynamics section.

### State transition probabilities

The agent-specific state transition probabilities depend on the age and precondition of the given agent. We consider seven age groups (0-4, 5-14, 15-29, 30-59, 60-69, 70-79, 80-) and four COVID-related chronic illnesses (besides the healthy) as preconditions: obstructive pulmonary, kidney and cardiovascular diseases, and diabetes in the model. The age group dependent epidemiological parameter values were published by Röst et al. (Table B3)[4]. Based on these average parameter values, we decreased the probability of the path towards the fatal outcome directly for the agents with healthy preconditions and modified the state transition probabilities of agents with chronic illness by an illness-specific multiplier. We increased the probability of death (uniformly on the path towards the deceased state, between the states) based on statistical data, which describes the mortality of infected people with different preconditions[6].

### Infection dynamics

For every ten-minute time-step, the number of agents at each location are counted. The weighted infectiousness of diseased agents is added up, further weighted by a location-specific factor for likelihood of transmission (1·0 indoors, 0·1 outdoors). The probability of infection is then calculated, and susceptible agents are infected based on this probability. It is defined by the following general formula: $P = 1 - e^{-\left(k\times l\times\frac{I}{N}\times t\right)}$, where $k$ is the disease-specific constant multiplied by a location-specific factor ($l$), $N$ is the number of total agents at the given place [7–9], $t$ is the exposure time (one time-step in our case) and $I = \sum_{k=1}^{i} Inf_k$ is the number of infectious agents weighted by their infectiousness ($Inf$). $k$ parameter was estimated by using Nelder-Mead method: we fitted the number of hospitalised agents to the normalised national data in the autumn wave.

### Theoretical considerations behind the proposed infection dynamics

The applied infection probability can be analytically derived using a local SEI type infection model as follows. The main modelling assumptions are: 1) a finite time interval of length $T$ is modelled in each simulation step, 2) the population is constant in the location during a time-step, 3) the number of infectious agents is constant during a time-step, 4) newly infected agents are transferred to the exposed ($E$) compartment, 5) all susceptible agents are in the $S$ compartment, 6) all infectious agents are modelled as members of the $I$ compartment.

Using these, the dynamical equations of the local infection model during a time interval $[0, T]$ can be written in the well-known kinetic form as

$$\dot{S} = -\frac{k}{N}\cdot I\cdot S,$$
$$\dot{E} = -\frac{k}{N}\cdot I\cdot S,$$
$$\dot{I} = 0 \rightarrow I = I(0) = const.$$

Using the assumptions, the above equations can be solved as

$$S(t) = S(0)e^{\left(-\frac{k}{N}\cdot I\cdot t\right)},$$

where $S(0)$ is the number of susceptible agents at the beginning of the time step. From this, we can integrate $E$ as well

$$E(t) = -S(t) + c = -S(0)e^{\left(-\frac{k}{N}\cdot I\cdot t\right)} + c,$$

where $c$ is a constant. According to our assumptions $E(0) = 0$ which implies $S(0) = c$ and therefore for the newly infected, we get

$$E(t) = S(0)\left(1 - e^{\left(-\frac{k}{N}\cdot I\cdot t\right)}\right)$$

Then the time-dependent infection probability can be computed as the relative frequency, i.e.

$$P(t) = \frac{E(t)}{S(0)} = 1 - e^{\left(-k\cdot\frac{I}{N}\cdot t\right)}$$

With this kind of modelling consideration and solution, a nice theoretical basis can be obtained for the good agreement between the stochastic and deterministic simulations using the fundamental results in the following paper of Thomas G Kurtz[10].

### COVID-19 unrelated health parameters

We also consider COVID-19 independent hospitalisation and death in the model, which is able to reconstruct the actual hospital burden observed in Szeged. Sex- and age-dependent data for mortality was obtained from the Hungarian Central Statistical Office, furthermore, quantitative information for hospitalisation probability, the average length of stay (5·55 days) at the hospital and mortality probability of hospitalised people was also available there. The sex- and age-dependency of hospitalisation and mortality rate were determined by using the data for average mortality. For agents with a special precondition, namely cardiovascular or obstructive pulmonary disease, unique hospitalisation probabilities and mortality probabilities of hospitalised agents were defined by using data from the Hungarian Central Statistical Office for the specific hospital wards. The probabilities can be found in Supplementary Table 5.

| Sex/Age | 0-4 | 5-14 | 15-29 | 30-59 | 60-69 | 70-79 | 80- |
|---|---|---|---|---|---|---|---|
| **Mortality probabilities outside of the hospital** | | | | | | | |
| **Male** | 3·80E-07 | 3·80E-07 | 3·80E-07 | 3·84E-06 | 2·01E-05 | 3·48E-05 | 1·05E-04 |
| **Female** | 2·03E-07 | 2·03E-07 | 2·03E-07 | 1·19E-06 | 8·87E-06 | 1·96E-05 | 9·74E-05 |
| **General hospitalisation probabilities** | | | | | | | |
| **Male** | 1·78E-04 | 1·19E-04 | 6·14E-04 | 3·17E-03 | 1·02E-03 | 1·10E-03 | 9·26E-04 |
| **Female** | 9·52E-05 | 6·35E-05 | 3·29E-04 | 9·87E-04 | 4·50E-04 | 6·24E-04 | 8·56E-04 |
| **General mortality probabilities of hospitalised agents** | | | | | | | |
| **Male** | 6·33E-03 | 4·22E-03 | 8·44E-03 | 3·02E-02 | 3·62E-02 | 3·92E-02 | 3·30E-02 |
| **Female** | 3·39E-03 | 2·26E-03 | 4·52E-03 | 9·39E-03 | 1·60E-02 | 2·22E-02 | 3·05E-02 |
| **Hospitalisation probabilities with cardiovascular disease** | | | | | | | |
| **Male** | 3·64E-04 | 2·43E-04 | 1·24E-03 | 6·39E-03 | 2·08E-03 | 2·26E-03 | 1·90E-03 |
| **Female** | 9·98E-05 | 6·65E-05 | 3·35E-04 | 9·99E-04 | 4·71E-04 | 6·54E-04 | 8·96E-04 |
| **Mortality probabilities of hospitalised agents with cardiovascular disease** | | | | | | | |
| **Male** | 2·75E-03 | 1·83E-03 | 3·67E-03 | 1·31E-02 | 1·57E-02 | 1·70E-02 | 1·43E-02 |
| **Female** | 1·47E-03 | 9·82E-04 | 1·96E-03 | 4·08E-03 | 6·96E-03 | 9·65E-03 | 1·32E-02 |
| **Hospitalisation probabilities with obstructive pulmonary disease** | | | | | | | |
| **Male** | 3·62E-04 | 2·41E-04 | 1·24E-03 | 6·38E-03 | 2·07E-03 | 2·24E-03 | 1·88E-03 |
| **Female** | 9·85E-05 | 6·57E-05 | 3·33E-04 | 9·96E-04 | 4·65E-04 | 6·45E-04 | 8·85E-04 |
| **Mortality probabilities of hospitalised agents with obstructive pulmonary disease** | | | | | | | |
| **Male** | 1·22E-02 | 8·14E-03 | 1·63E-02 | 5·83E-02 | 6·98E-02 | 7·57E-02 | 6·36E-02 |
| **Female** | 6·54E-03 | 4·36E-03 | 8·72E-03 | 1·81E-02 | 3·09E-02 | 4·28E-02 | 5·87E-02 |

**Supplementary Table 5: COVID-19 unrelated health parameters.** The table summarises the hospitalisation and mortality rates of people with certain chronic diseases. The Szeged specific values were derived from data from the Hungarian Central Statistical Office.

### Fitting the parameters to real life data from Hungary and to a deterministic SEIRD model

To fine-tune the parameters of the agent-based model (time constants, transition probabilities), we have used the validated and published deterministic model[4,5] as well. We have selected the autumn wave of the epidemic to fit the model to actual data. The output (measured) variables were the daily total and positive tests, the daily number of hospitalised people and the cumulative number of deaths which were assumed to be reliable enough for fitting the model since it is known that the number of officially recorded daily infections is only a fraction of the true cases. Most importantly, we have fitted the number of hospitalised COVID-19 patients to the national data scaled with the ratio of populations by setting the disease-specific infectivity parameter. As a result, the number of hospitalised, deceased people due to COVID-19 and daily testing rate and the ratio of positive tests match well to the real data (Supplementary Figure 4).

We have also used the special mathematical property of the deterministic model[5] that the number of latent infected people can be computed by the dynamic inversion of a linear subsystem of the compartmental model. Then it is possible to estimate the non-measured internal state variables (exposed, presymptomatic, asymptomatic, symptomatic infected, recovered, susceptible) using a standard state estimator with guaranteed stability and convergence. From these data, we can also compute acceptable estimates for the probable true number of daily infections, the number of susceptible people (taking into consideration the vaccination data in the later stage of the epidemic) and to the time-dependent, effective reproduction number ($R_t$). The simulations of the agent-based model also match to the data obtained from the applied deterministic model. The non-measured internal state variables in the two simulations give a satisfactory fit, too (Supplementary Figure 5).

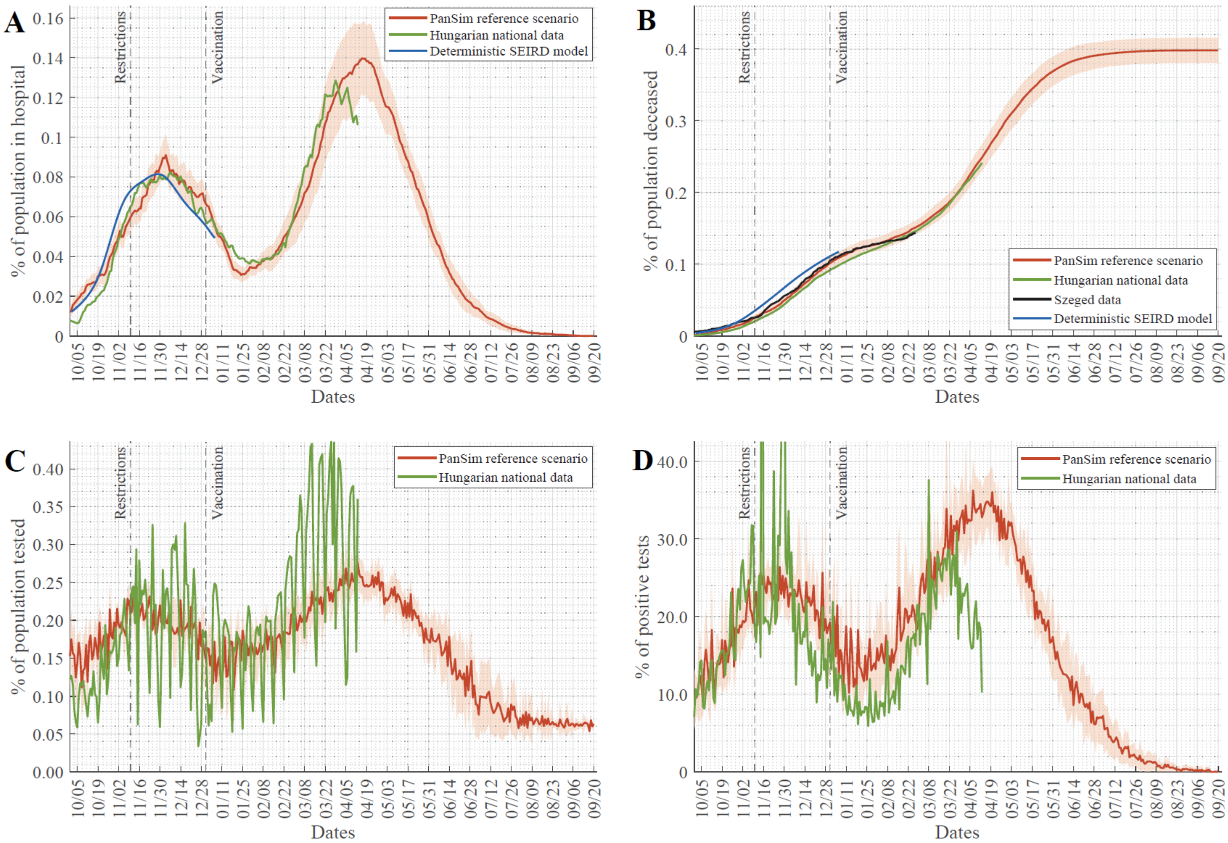


**Supplementary Figure 4:** Fitting the parameters of a deterministic SEIRD model and *PanSim* to real data. (A) Percentage of hospitalised COVID-19 patients in the population, (B) accumulated number of death events due to COVID-19 scaled to the whole population, (C) daily testing rate and (D) daily ratio of positive tests were collected for the city of Szeged (black) or calculated by population size ratio of Szeged from Hungarian national data (green). A deterministic SEIRD model[5] was fitted to this data (blue) for the whole length of the autumn wave (until 1st January 2021). The SEIRD model was used to predict changes in SEIR measures (Supplementary Figure 5.), and all these data were used to fit parameters of *PanSim* (red – average and std. of 20 simulations).

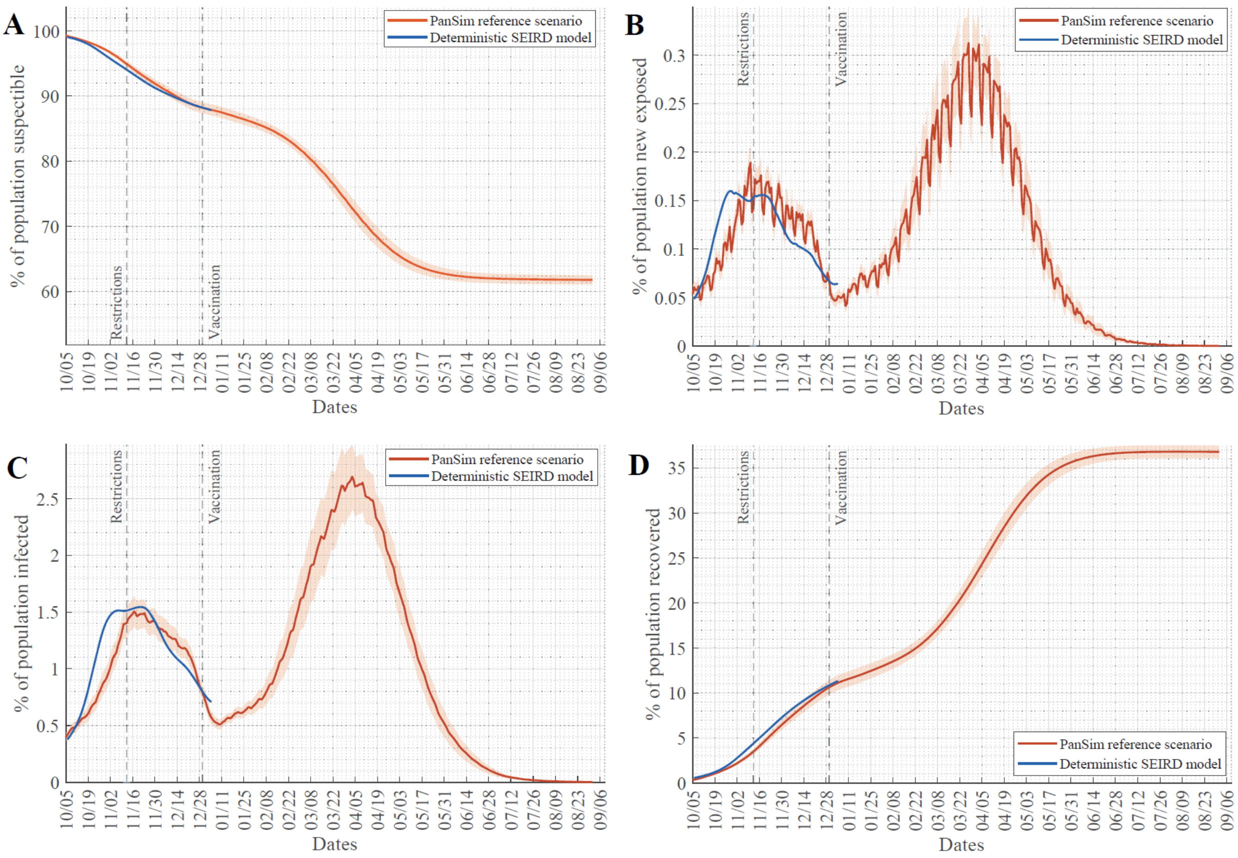


**Supplementary Figure 5:** Fitting the parameters of *PanSim* to SEIR measure predictions of a deterministic model. Since there is no precise empirical data on the percentage of susceptible (A), new COVID-19 exposed (B), infected (C) and recovered (D) individuals in the population of Szeged, we have simulated these values for the autumn wave from the deterministic model (blue) that was fitted to real data (Supplementary Figure 4.). These plots were matched by the fitted *PanSim* model (red - average and std. of 20 simulations).

### Simulator functionalities

To realistically simulate the spread of infection, as well as the effect of countermeasures, we have designed and developed an agent-based simulator. It creates virtual agents, each with its own properties such as age, sex, preconditions, current disease-related state, and a list of specific locations, such as the home, workplace, preferred grocery stores, etc. associated with the agent, as well as an agent profile (such as full-time worker, or retired, etc.) which determines its movement patterns. At the same time, virtual locations are created, each with its own type (school, restaurant, residency, etc.) – these hold a list of agents present at that location.

#### Movement

The simulator is driven on a daily loop: at midnight, statistics are calculated, the disease progression is updated for each agent, restrictions are enabled or disabled for various locations, and a variety of predefined events are triggered. Then, during the day, agents can move between locations and infect each other at a time resolution of 10 minutes by default. The movement of agents is determined by their schedule using time windows: in any given time window, an agent will randomly choose a target location type, weighted by probabilities given in the schedule for that window, then randomly pick a specific location that is associated with that agent and has the correct type. The agent will then spend a predetermined amount of time there. For example, an adult agent between 5 pm and 6 pm will leave for home with 70% probability and stay there for two hours, or leave for a small social gathering place with 30% probability and stay there for 1·5 hours.

#### Restrictions

An important factor affecting the movement of agents is the closing of certain types of locations. These can either be triggered on particular days or when a certain threshold is met (e.g. number of people in hospitals above a level for four consecutive days). When an agent tries to go to a closed location, it will try to choose a different type of location (according to its schedule), and if still, none is available, it will go home. There are restriction events that apply to all locations of a particular type (e.g. restaurants), or that limit children of a certain age going to school (e.g. no school for age 12 and above), or that impose a curfew in a given part of the day, at which time only those agents with a non-standard work schedule can go to work.

#### Testing schemes

Testing agents is a dominant element of controlling the pandemic. In our simulator, agents who feel ill with the disease and go to a doctor's office or to a hospital get automatically diagnosed. Diagnosis can also be triggered by ways of randomised testing, for which we have multiple categories. The probability of any given agent getting tested on a particular day depends on an overall random probability, whether someone has been diagnosed in the last 24 hours who lives in the same household or goes to the same classroom or workplace, and whether the agent works in a public health institute or lives/works in a nursing home. Agents can get tested up to every 5 days by default. The sensitivity of testing can be further specified to allow differentiation between PCR and antibody tests. The following testing probabilities were set in the reference scenario:

- random: 0.005%,
- home: 1%,
- workplace: 0.05%,
- class: 0.05%,
- hospital worker: 0.5%,
- nursing home: 5%

The following parameters were modified in the scenario, where the testing probabilities of contacts were increased (Figure 2C-D):

- home: 100%,
- workplace: 20%,
- class: 20%

#### Quarantining measures

Another important factor in curbing the spread of the disease is quarantining agents. When an agent is diagnosed, depending on a quarantine level setting, the following will happen:

- Level Q0: the agent is not quarantined
- Level Q1: only the diagnosed agent is quarantined
- Level Q2: the diagnosed agent and everyone else in the same household are quarantined
- Level Q3: the agent, household, and everyone in the same classroom or a portion of workmates is quarantined
- Level Q4: as with Level Q3, but when at least 3 classrooms in the same school get quarantined, the whole school gets quarantined

An agent that is in quarantine will not leave its home for a certain amount of time (10 days by default) unless they are taken to the hospital.

#### Virus variants

As the B.1.1.7 variant now accounts for most infections in Szeged, a new strain was introduced to the model, which has different (1·66 times higher) infectiousness and distinct disease progression properties (the parameter values describing the probability of hospitalisation and the probability of fatal outcome are 1·2 times higher)[11]. In the simulator, the two are handled separately in terms of infection events but share the same SEIRD states, albeit with modified transition probabilities - recovered agents do not get infected with either strain.

The new variant can be introduced into the population with a predefined event that will randomly infect a given number of susceptible agents at a given time.

#### Vaccination

After a given time into the simulation, immunisation can be enabled at a given rate - or even at variable rates to account for real vaccination statistics or predictions. Agents to be vaccinated are grouped by a number of properties (age, preconditions, occupation, etc.) and these groups are immunised in a given order - each group at a given level (depending on willingness). After the vaccination event, an agent's probability of being infected will be reduced at a predefined rate, currently set to 52% after 12 days and 96% after 28 days[12].
The following vaccine prioritisation rules were analysed with the model:
Vaccination order based on vulnerability risk:

1. 60+ with an underlying health condition
2. 60+
3. 18-60 with an underlying health condition
4. 18+

Vaccination order based on occupational risk:

1. health workers
2. nursery home workers and residents
3. essential workers
4. school teachers
5. 18+

Mixed vaccination order (mimicking the Hungarian vaccination plan):

1. health workers
2. nursery home workers and residents
3. 60+
4. 18-60 with an underlying health condition
5. essential workers
6. 18+

### Parameter sets

#### Reference scenario

The results in Figure 1C were derived from our "reference scenario" that takes the following measures and parameters into consideration:

- testing probabilities:
    - random: 0·005%,
    - home: 1%,
    - workplace: 0·05%,
    - class: 0·05%,
    - hospital worker: 0·5%,
    - nursing home: 5%
- Restrictions
    - from 1st October, closed:
        - nursing homes,
        - entertainment sites
    - from 11th November, closed:
        - high schools and universities,
        - evening-schedule services
        - a curfew between 8 pm – 5 am
- no reopening

– household of those diagnosed are automatically quarantined for 10 days, for children so are classmates, and for adults working in offices, colleagues are quarantined as well (Level Q3)
– vaccination starts on 1st January, with 0·2% of the population vaccinated daily based on the “Mixed” vaccination order

The rest of the discussed simulations use the same parameters unless otherwise stated.

#### “Unmitigated” scenario (used in Figure 1B)

The unmitigated scenario has the same parameters as the reference scenario with the differences that intervention measures are not applied. There are no restrictions, curfew and quarantine, and all locations are open.

| Parameter | Scenarios | Total number of deaths (D) | Sum of the hospital capacity overrun (H) | Severity function |
|---|---|---|---|---|
| | | mean (+/-standard deviation) | | standardized values |
| | Reference scenario | 714·5 (+/-31·37) | 1545·4 (+/-1097·18) | 0 |
| Closed from 11th November | Large events, nursing homes | 869·6 (+/-45·98) | 3363·95 (+/-1665·20) | 0·5332906586 |
| | All non-essential locations, evening curfew | 415·95 (+/-27·39) | 0 | -0·9830370331 |
| Quarantined people | Q1: diagnosed only | 748·05 (+/-28·09) | 9289·7 (+/-855·13) | 0·278863632 |
| | Q2: Q1 + household of diagnosed | 794·05 (+/-35·04) | 2285·25 (+/-1289·07) | 0·2692319486 |
| Testing probability | 10 times higher | 654·5 (+/-41·21) | 1803·05 (+/-1188·03) | -0·1849231418 |
| | 20 times higher | 637·9 (+/-40·02) | 2116·5 (+/-1453·09) | -0·230698701 |
| Reopening time | 1st April | 922·95 (+/-40·01) | 7818·1 (+/-1625·20) | 0·8018874115 |
| | 1st May | 803·5 (+/-41·10) | 1066·65 (+/-850·91) | 0·2721546751 |
| | 1st June | 728 (+/-27·92) | 1109·95 (+/-977·87) | 0·03321145774 |
| Infectivity of novel variant | 1·9 times higher | 959 (+/-28·53) | 11574·85 (+/-1538·9) | 1 |
| | 1·5 times higher | 534 (+/-43·28) | 0 | -0·6079252026 |
| Vaccinated daily | 0·1% | 926·75 (+/-43·21) | 6128·85 (+/-1827·50) | 0·7763882276 |
| | 0·3% | 543·2 (+/-39·76) | 73 (+/-292·25) | -0·5770678457 |
| Vaccination order by | Vulnerability | 640·65 (+/-27·98) | 1041·35 (+/-1108·53) | -0·2458749217 |
| | Occupational risk | 785·65 (+/-29·05) | 3222·4 (+/-974·19) | 0·2633853835 |

**Supplementary Table 6:** Table of the severity function. The table contains the results of scenarios in Figure 1C. The mean and standard deviation values of the total number of deaths (D) and the sum of hospital capacity overrun (H) were calculated from 20 simulations. The standardised values of the *severity function* (S) are calculated as $S = \frac{(D+0.007H)-S_{Reference\ scenario}}{S_{max}}$ and it is characterised by colour code.

### Implementation

The code is implemented in the C++ language, using the Thrust library to target multi-core CPU and GPU architectures. Particular care was given to efficiency – we can currently simulate Szeged for one year with a ten minute time-step and 179500 agents in 57 seconds on a single NVIDIA V100 GPU, and the code is scalable up to 90 million agents on a single GPU. Our full simulator code is available at https://github.com/khbence/pansim, completed with input files and default parameter settings used for simulations in this paper.

#### Output reporting

The PanSim produced different types of outputs:

1. Standard, time-series data (one txt file per run): time-series of the compartments of the SEIRD-model, number of tests, quarantine, vaccination, the ratio of novel virus variant.
2. Agent list (one JSON file per run): list of agents, who got infected at a given time and place. The time of diagnosis, duration of infection, quarantine, hospitalisation and worst state, the time of immunisation is also given.
3. List of agent movements (one txt file per 10-minute time-step in the simulation): listing where each person is and their infection status.
4. List of infection events (one txt file per 10-minute time-step in the simulation): all of the infection events occurred in a given time-step, with different locations, people got infected there, and people that infected them.

These outputs are processed by MATLAB scripts: separate parts are responsible for processing one of the main inputs. The code is able to process any given number of different scenarios, and in the case of each and every scenario, an arbitrary number of stochastic simulations can be used for computing average and standard deviation.

The MATLAB scripts give back different types of output by using the results of the PanSim as input:

1. Visualisation of the standard, time-series data: the mean time-series with the standard deviation is computed for every scenario, which are plotted together. A time-dependent reproduction number ($R_t$) is also computed by taking the total new cases into account. We consider a 10-day window as an average duration for being infected: duration of 9·5 days is the average period without hospitalisation, duration of 19·5 days is the average period with hospitalisation, and we assumed 5% probability for being hospitalised, therefore by taking a weighted average we get 10 days as an average duration. At each timepoint, $R_t$ refers to the ratio of the total number of new cases in the current day and next 9 days and in the previous 10 days. Another more direct, contact tracing-based approach is being developed as an agent-level alternative way for calculating $R_t$: the simulation can track who infected whom (with some probabilities if multiple infectious agents are present at the same location).
2. A CSV file of a few simple metrics: a CSV file is generated, which is a 12×N table, where N is the number of examined scenarios. The first column is the scenario ID in the form of a string and is followed by the mean and standard deviation of the metrics. The metrics are the following: total number of the deceased people, the total number of hospital bed days, maximum number of people in hospital recorded, total number of intensive care bed days, maximum number of people in intensive care recorded, the total number of hospital beds occupied above a critical limit.
3. Visualisation of agent list data: several runs are processed, statistics are prepared by computing means and standard deviations and plotting them in the form of bar plots. We can measure the distributions of infected, hospitalised population or deceased people according to age, precondition, sex, worst state, agent type and location type of infection.
4. Visualisation of agent movements: a separate Matlab script reads agent location data and renders a video that visualises the spatial distribution of the total number of agents or the number of infected agents in time. The visualisation is aligned on an actual map of the simulated area, and the different types of locations are colour-coded. The progress of the simulation and the dynamics of the actual number of the hospitalised are easy to follow.

### References


1 COVID-19 Community Mobility Reports. https://www.google.com/covid19/mobility/?hl=en-GB (accessed 11th May, 2021).

2 Buckee CO, Balsari S, Chan J, *et al.* Aggregated mobility data could help fight COVID-19. Science (80-. ). 2020; **368**: 145–6.

3 Szocska M, Pollner P, Schiszler I, *et al.* Countrywide population movement monitoring using mobile devices generated (big) data during the COVID-19 crisis. *Sci Rep* 2021; **11**: 5943.

4 Röst G, Bartha FA, Bogya N, *et al.* Early phase of the COVID-19 outbreak in Hungary and post-lockdown scenarios. *Viruses* 2020; **12**: 1–30.

5 Péni T, Csutak B, Szederkényi G, Röst G. Nonlinear model predictive control with logic constraints for COVID-19 management. *Nonlinear Dyn* 2020; **102**: 1965–86.

6 Banerjee A, Pasea L, Harris S, *et al.* Estimating excess 1-year mortality associated with the COVID-19 pandemic according to underlying conditions and age: a population-based cohort study. *Lancet* 2020; **395**: 1715–25.

7 Balcan D, Gonçalves B, Hu H, Ramasco JJ, Colizza V, Vespignani A. Modeling the spatial spread of infectious diseases: The global epidemic and mobility computational model. *J Comput Sci* 2010; **1**: 132–45.

8 Chinazzi M, Davis JT, Ajelli M, *et al.* The effect of travel restrictions on the spread of the 2019 novel coronavirus (COVID-19) outbreak. *Science (80- )* 2020; **368**: 395–400.

9 Chang S, Pierson E, Koh PW, *et al.* Mobility network models of COVID-19 explain inequities and inform reopening. *Nature* 2021; **589**: 82–7.

10 Kurtz TG. The relationship between stochastic and deterministic models for chemical reactions. *J Chem Phys* 1972; **57**: 2976–8.

11 Karin O, Bar-On YM, Milo T, *et al.* Cyclic exit strategies to suppress COVID-19 and allow economic activity. medRxiv. 2020; : 2020.04.04.20053579.

12 Polack FP, Thomas SJ, Kitchin N, *et al.* Safety and Efficacy of the BNT162b2 mRNA Covid-19 Vaccine. *N Engl J Med* 2020; **383**: 2603–15.
